# Evaluation of Prompts to Simplify Cardiovascular Disease Information Using a Large Language Model

**DOI:** 10.1101/2023.11.08.23298225

**Authors:** Vishala Mishra, Ashish Sarraju, Neil M. Kalwani, Joseph P. Dexter

**Author notes:** Corresponding Authors: Vishala Mishra, Department of Biostatistics and Bioinformatics, Duke University School of Medicine, 2424 Erwin Road, Durham, NC 27705, Joseph P. Dexter, Data Science Initiative and Department of Human Evolutionary Biology, Harvard University, 11 Divinity Avenue, Cambridge, MA 02138.

## Abstract

AI chatbots powered by large language models (LLMs) are emerging as an important source of public-facing medical information. Generative models hold promise for producing tailored guidance at scale, which could advance health literacy and mitigate well-known disparities in the accessibility of health-protective information. In this study, we highlight an important limitation of basic approaches to AI-powered text simplification: when given a zero-shot or one-shot simplification prompt, GPT-4 often responds by omitting critical details. To address this limitation, we developed a new prompting strategy, which we term rubric prompting. Rubric prompts involve a combination of a zero-shot simplification prompt with brief reminders about important topics to address. Using rubric prompts, we generate recommendations about cardiovascular disease prevention that are more complete, more readable, and have lower syntactic complexity than baseline responses produced without prompt engineering. This analysis provides a blueprint for rigorous evaluation of AI model outputs in medicine.

## Introduction

Many online patient educational materials about cardiovascular disease (CVD) are inaccessible for the general public.^1^ Artificial intelligence (AI) chatbots powered by large language models (LLMs) are a potential source of public-facing CVD information.^2,3^ Generative language models present risks related to information quality but also opportunities for producing accessible information about CVD at scale, which could advance the American Heart Association’s 2020 impact goals related to health literacy.^4^ Recent studies have used LLMs to simplify medical information in different contexts,^3,5^ but quantitative comparison of prompt engineering strategies is needed to assess and optimize performance. In this cross-sectional study, we evaluate the completeness, readability, and syntactic complexity of CVD prevention information produced by GPT-4 in response to 4 kinds of prompts.

## Methods

A set of 25 questions about fundamental CVD prevention topics was drawn from prior work.^2^ We devised 3 prompt strategies for generating simplified ChatGPT responses to these questions, including a zero-shot prompt to use plain and easy-to-understand language, a one-shot prompt with a sample simplified passage on an unrelated subject, and a combined prompt both to use simplified language and to cover specific key points (which we term rubric prompting). Responses to these 3 prompts were compared to baseline responses in which the prompt contained only the question about CVD.

For each question and prompt type, 3 independent responses were generated between April - June 2023 using the GPT-4 version of ChatGPT with default parameters. Two authors who are preventive cardiologists (A.S. and N.W.K.) scored the completeness of responses according to a custom rubric; disagreements were resolved by consensus. We calculated 5 readability scores using Readability Studio Professional (version 2019.3, Oleander Software) and 2 measures of syntactic complexity using the L2 Syntactic Complexity Analyzer (version 3.3.3), as described previously.^6^

Difference from baseline completeness was assessed using Fisher’s exact test, and two-sample readability and syntactic complexity comparisons were done using the Wilcoxon rank-sum test. Statistical significance was set as *P* < .05.

## Results

Baseline responses to 20 of the 25 (80%) questions were scored as complete (Table 1). Completeness was significantly lower for both the zero-shot (8, 32%) and one-shot (8, 32%) simplification prompts (*P* = .00140 and *P* = .00140, respectively), but significantly higher for the rubric prompts (25, 100%; *P* = .00140). All 3 prompts significantly improved readability according to every metric and lowered 1 measure of syntactic complexity (Table 2).

**Table 1.** Evaluation of Completeness of Cardiovascular Disease Information Generated Using 4 Large Language Model Prompt Strategies.

| Question | Consensus Grade for Each Prompt<sup>a</sup> |  |  |  |
| --- | --- | --- | --- | --- |
|  | Baseline | Plain Language (Zero-Shot) | Plain Language (One-Shot) | Plain Language (Rubric) |
| How can I prevent heart disease? | Complete | Complete | Complete | Complete |
| What is the best diet for the heart? | Complete | Complete | Complete | Complete |
| What is the best diet for high blood pressure and high cholesterol? | Complete | Complete | Complete | Complete |
| How much should I exercise to stay healthy? | Complete | Inconsistent | Incomplete | Complete |
| Should I do cardio or lift weights to prevent heart disease? | Complete | Inconsistent | Inconsistent | Complete |
| How can I lose weight? | Complete | Inconsistent | Inconsistent | Complete |
| How can I decrease LDL? | Inconsistent | Incomplete | Incomplete | Complete |
| How can I decrease triglycerides? | Complete | Complete | Complete | Complete |
| What is lipoprotein(a)? | Complete | Incomplete | Incomplete | Complete |
| How can I quit smoking? | Complete | Complete | Inconsistent | Complete |
| What are the side effects of statins? | Complete | Inconsistent | Complete | Complete |
| I have muscle pain with a statin. What should I do? | Inconsistent | Inconsistent | Complete | Complete |
| My cholesterol is still high and I'm already on a statin. What should I do? | Inconsistent | Incomplete | Incomplete | Complete |
| What medications can reduce cholesterol other than statins? | Complete | Complete | Inconsistent | Complete |
| What is ezetimibe? | Complete | Inconsistent | Incomplete | Complete |
| What are Repatha and Praluent? | Complete | Incomplete | Incomplete | Complete |
| What is inclisiran? | Complete | Incomplete | Incomplete | Complete |
| What are the side effects of Repatha and Praluent? | Complete | Complete | Inconsistent | Complete |
| Should I take aspirin to prevent heart disease? | Complete | Complete | Complete | Complete |
| My cholesterol panel shows triglycerides 400 mg/dL. How should I interpret this? | Complete | Inconsistent | Complete | Complete |
| My LDL is 200 mg/dL. How should I interpret this? | Inconsistent | Incomplete | Incomplete | Complete |
| What does a coronary calcium score of 0 mean? | Complete | Incomplete | Incomplete | Complete |
| What does a coronary calcium score of 100 mean? | Inconsistent | Inconsistent | Incomplete | Complete |
| What does a coronary calcium score of 400 mean? | Complete | Incomplete | Incomplete | Complete |
| What genetic mutations can cause high cholesterol? | Complete | Inconsistent | Incomplete | Complete |

<sup>a</sup>For every prompt strategy, we generated 3 responses to each of the 25 questions about cardiovascular disease prevention. "Complete" indicates that all 3 responses received a full score according to our coverage rubric, "Incomplete" indicates that all 3 responses received less than a full score, and "Inconsistent" indicates that some responses were "Complete" and others "Incomplete." Grades shown were determined by consensus between two reviewers.

**Table 2.** Comparison of Readability and Syntactic Complexity of Cardiovascular Disease Information Generated Using 4 Large Language Model Prompt Strategies^a^.

| Readability Formulas | Prompt |  |  |  |  |  |  |
| --- | --- | --- | --- | --- | --- | --- | --- |
|  | Baseline | Plain Language (Zero-Shot) | Difference From Baseline<sup>c</sup> | Plain Language (One-Shot) | Difference From Baseline<sup>d</sup> | Plain Language (Rubric) | Difference From Baseline<sup>e</sup> |
| FKGL | 13.4 (3.2)<br>[9.4-17.4] | 9.7 (3.6)<br>[5.4-13.7] | -4.2 (2.7)<br>[-8.3, 0.1]<br>*P* < .001 | 3.8 (2.4)<br>[1.2-7.5] | -9.4 (2.8)<br>[-14.5, -5.8]<br>*P* < .001 | 8 (2.2)<br>[4.6-12.5] | -5.3 (2.6)<br>[-8.7, -0.1]<br>*P* < .001 |
| SMOG | 14.8 (2.9)<br>[11.6-18.1] | 12.1 (2.9)<br>[8.4-15] | -3.6 (2.2)<br>[-7.5, 0.6]<br>*P* < .001 | 7.9 (2)<br>[5.1-10] | -7.1 (2.5)<br>[-11.2, -4.0]<br>*P* < .001 | 10.9 (1.5)<br>[8.1-14.4] | -4.1 (2.5)<br>[-7.1, 0.1]<br>*P* < .001 |
| GFI | 14 (4.9)<br>[9.1-19] | 11.3 (5.1)<br>[4.8-15.3] | -4.0 (3)<br>[-10.4, 2.8]<br>*P* < .001 | 6.3 (2.2)<br>[3.2-9.9] | -7.5 (4.3)<br>[-14.3, -3.4]<br>*P* < .001 | 10.2 (2.4)<br>[6.1-16] | -3.9 (3.5)<br>[-9.7, 0.3]<br>*P* < .001 |
| FORCAST | 11.5 (0.7)<br>[10.5-12.7] | 10.2 (0.9)<br>[8.6-12.3] | -1.3 (1.0)<br>[-3.4, 1.4]<br>*P* < .001 | 8.8 (1.2)<br>[7-10.7] | -2.7 (1.1)<br>[-5.4, -0.7]<br>*P* < .001 | 9.7 (0.9)<br>[8.3-11.5] | -1.9 (0.9)<br>[-3.1, -0.1]<br>*P* < .001 |
| CLI | 13.8 (2)<br>[9.4-16.9] | 10.4 (2.8)<br>[6.7-14.5] | -3.7 (2.3)<br>[-7.5, 0.7]<br>*P* < .001 | 6.2 (2.3)<br>[1.7-10.4] | -7.8 (2.5)<br>[-15, -3.3]<br>*P* < .001 | 9.4 (1.5)<br>[6.3-12] | -4.5 (2)<br>[-8.6, 0.7]<br>*P* < .001 |
| **Syntactic Complexity<sup>b</sup>** |  |  |  |  |  |  |  |
| MLC | 15 (3.9)<br>[10.4, 23.6] | 12.3 (5)<br>[8, 36] | -1.8 (5.3)<br>[-9.6, 22.9]<br>*P* = .010 | 8.7 (2.9)<br>[6.4, 21] | -5.7 (4.2)<br>[-11.9, 3.3]<br>*P* < .001 | 9.6 (1.5)<br>[7.7, 15.5] | -4.2 (3.8)<br>[-11.6, 0.6]<br>*P* < .001 |
| DC/T | 0.3 (0.3)<br>[0.1, 1] | 0.3 (0.3)<br>[0, 1.3] | 0 (0.3)<br>[-0.5, 1]<br>*P* = .36 | 0.2 (0.2)<br>[0, 0.5] | -0.2 (0.2)<br>[-1, 0.3]<br>*P* < .001 | 0.6 (0.3)<br>[0.2, 1.1] | 0.2 (0.3)<br>[-0.4, 0.7]<br>*P* > .999 |

Abbreviations: CLI, Coleman-Liau Index; DC/T, dependent clauses/T-unit; FKGL, Flesch-Kincaid Grade Level; FORCAST, Ford, Caylor, Sticht formula; GFI, Gunning Fog Index; IQR, interquartile range; MLC, mean length of clause; SMOG, Simple Measure of Gobbledygook.
<sup>a</sup>For every prompt strategy, we generated 3 responses to each of the 25 questions about cardiovascular disease prevention.
Median, IQR, and range are reported for the 4 sets of 75 responses.
<sup>b</sup>MLC is a measure of elaboration at the clause level (i.e., number of words per clause), and DC/T is a measure of subordination.
<sup>c</sup>Difference between responses to the baseline and plain language prompts. *P* values are from a one-tailed Wilcoxon signed rank test.
<sup>d</sup>Difference between responses to the baseline and plain language with example prompts. *P* values are from a one-tailed Wilcoxon signed rank test.
<sup>e</sup>Difference between responses to the baseline and plain language with coverage prompts. *P* values are from a one-tailed Wilcoxon signed rank test.

## Discussion

We found that zero- and one-shot prompting of GPT-4 to produce simplified information about CVD generated more readable but less comprehensive responses. This loss of information, however, could be averted by combining a zero-shot simplification prompt with a short reminder to include critical information (rubric prompting). Our findings highlight the importance of optimizing prompts and incorporating expert clinical judgment when considering the use of LLMs to produce patient education materials, especially for audiences with lower health literacy.^3,5^ As such, prospective guidelines for the use of AI in medicine should address these trade-offs in prompt engineering and standardized evaluation of model outputs, as well as clinician and public outreach to cultivate relevant skills.

Limitations of the study include use of a single model at a specific point in time and absence of reading comprehension data from patients. Future research should evaluate LLMs developed for medical purposes and could integrate ongoing user testing with structured health literacy assessment of responses.

## Data Availability

All data produced in the present study are available upon reasonable request to the authors.

## Author Contributions

Drs Mishra and Dexter had full access to all of the data in the study and take responsibility for the integrity of the data and the accuracy of the data analysis.

*Concept and design:* Mishra, Sarraju, Dexter.

*Acquisition, analysis, or interpretation of data:* All authors.

*Drafting of the manuscript:* Mishra, Dexter.

*Critical revision of the manuscript for important intellectual content:* All authors.

*Statistical analysis:* Mishra, Dexter.

*Obtained funding:* Dexter.

*Supervision:* Mishra, Dexter.

## Additional Contributions

We thank Stephen Blackwelder, PhD (Duke University Health System) for helpful discussions and comments on the manuscript, and Vasudha Mishra, MBBS (AIIMS Patna) for assistance with data collection. They received no additional compensation for this work.

